# Two-sample Mendelian randomization analysis of associations between periodontal disease and risk of colorectal, lung, and pancreatic cancers

**DOI:** 10.1101/2021.01.14.21249587

**Authors:** Laura Corlin, Mengyuan Ruan, Konstantinos K. Tsilidis, Emmanouil Bouras, Yau-Hua Yu, Rachael Stolzenberg-Solomon, Alison P. Klein, Harvey A. Risch, Christopher I. Amos, Lori C. Sakoda, Pavel Vodička, Rish K. Pai, James Beck, Elizabeth A. Platz, Dominique S. Michaud

## Abstract

Observational studies indicate that periodontal disease may increase the risk of colorectal, lung, and pancreatic cancers. We tested these associations using two-sample Mendelian randomization to emulate a randomized study with observational data. We developed an instrument including single nucleotide polymorphisms with strong genome-wide association study evidence for associations with aggressive and/or advanced periodontal disease. We used this instrument to assess associations with summary-level genetic data for colorectal cancer (n=58,131 cases), lung cancer (n=18,082 cases), and pancreatic cancer (n=9254 cases). The genetic predisposition index for periodontitis was significantly associated with an increased risk of colorectal cancer (p=0.026), colon cancer (p=0.021), proximal colon cancer (p=0.013), and colorectal cancer among females (p=0.039); however, it was not significantly associated with the risk of lung cancer or pancreatic cancer, overall or within most subgroups. Further research should determine whether increased periodontitis prevention and increased cancer surveillance of patients with periodontitis is warranted.

## Introduction

Colorectal cancer, lung cancer (including tracheal and bronchus cancers), and pancreatic cancer together account for over 2.9 million deaths per year globally.^1^ Colorectal cancer and lung cancer have the two highest number of incident cases globally (1.7 million and 2.0 million cases per year, respectively). Most histologic sub-types of lung cancer and pancreatic cancer have particularly poor prognoses since, even in the United States, at least 50% of cases are not diagnosed until the cancer is at a less curative, advanced stage.^2^ Primary prevention efforts are thus critical, and observational studies indicate that modifiable risk factors (e.g., periodontal disease), are implicated in the pathogenesis of colorectal, lung, and pancreatic cancers.^3–5^ Although well-conducted meta-analyses of these and other observational studies have strengthened the evidence for positive associations between periodontal disease and lung, colorectal, and pancreatic cancers,^6–8^ additional evidence for causal associations could be observed in randomized trials or in observational studies that employ methods that emulate randomization (e.g., Mendelian randomization; MR).

Two-sample MR is an approach that uses summary association estimates (often from genome-wide association studies; GWAS) to develop a genetic instrument index for the exposure, and then applies the index to assess the association with the outcome in a different sample of the same underlying source population.^9,10^ The instrument must be associated with the exposure, associated with the outcome only through paths that include the exposure, and independent of exposure-outcome confounders.^11^ Two-sample MR has advantages over one-sample MR: weak instrument bias tends to drive association estimates towards the null in two-sample MR rather than in the direction of the observational associations as in one-sample MR, and robust instruments from larger GWAS can be used in two-sample MR investigations such that more precise and accurate estimates may be obtained.^12–14^

Previous two-sample MR studies used genetic instruments for periodontal disease^15–17^ and other studies used two-sample MR to assess risk factors for each of colorectal, lung, and pancreatic cancer;^18–20^ however, to our knowledge, no MR study has assessed the association between periodontal disease and cancer risk. Given the need to rigorously assess putative causal relationships among modifiable factors (e.g., oral health) and cancer risk to support health promotion, our primary objective was to assess whether genetic predisposition to having chronic or aggressive periodontal disease was associated with colorectal, lung, or pancreatic cancers using a two-sample MR analysis. Our secondary objectives were to assess whether these associations (1) varied by cancer sub-type (location in the large bowel for colorectal cancer and histology for lung cancer), sex (for colorectal and pancreatic cancers), or smoking history (for lung and pancreatic cancers); and (2) were robust against potential violations of MR assumptions.

## Results

Using the eight SNPs with the strongest evidence for a genetic predisposition to having chronic or aggressive periodontal disease (Table S1), we observed a statistically significant association with the risk of colorectal cancer (3% increase per unit increase in genetic index of periodontal disease; p = 0.026) but not with the risk of lung (0.4% increase; p = 0.832) or pancreatic cancers (2% increase; p = 0.506; Table 1 and Figure 1). In secondary analyses, including an additional six SNPs with moderately strong evidence for an association with periodontitis attenuated the effect estimates for the association with colorectal cancer but did not substantially change the effect estimates for either lung or pancreatic cancers. For the primary and secondary analyses, *I*^2^_*GX*_ values were 0.947 and 0.926, respectively, indicating that the effect estimates were unlikely to be substantially biased towards the null due to violations of the NOME assumption. There was no indication of potential horizontal pleiotropy for any of the primary analyses (*I*^2^ = 0% for each colorectal, lung, and pancreatic cancers) and limited evidence of heterogeneity for the secondary analyses (*I*^2^ = 60% for colorectal, 12% for lung, and 0% for pancreatic cancer). Separate sensitivity analyses removing each of (1) rs1537415 (palindromic allele), (2) rs3826782 (low effect allele frequency and potentially influential), (3) rs12461706 (palindromic allele), (4) rs1537415 and rs12461706 (palindromic alleles in the primary instrument), and (5) rs9984417 (palindromic allele) did not substantively change any of these results.

**Table 1.** Effect estimates for the association between genetic predisposition to having chronic or aggressive periodontitis and the risk of colorectal, lung, and pancreatic cancer by genetic instrument and MR approach.

| Cancer outcome | N<sub>cases</sub> / N<sub>controls</sub> | Instrument<sup>1</sup> | IVW<sup>2</sup><br>β (p-value) | MR-PRESSO<br>β (p-value) | Simple median<br>β (p-value) | Weighted median<br>β (p-value) |
| --- | --- | --- | --- | --- | --- | --- |
| Colorectal | 58131/67347 | Primary | 0.025 (0.026) | 0.025 (0.010) | 0.025 (0.115) | 0.027 (0.063) |
|  |  | Secondary | 0.006 (0.699) | 0.016 (0.106) | 0.002 (0.875) | 0.025 (0.054) |
| Colon | 31083/67347 | Primary | 0.031 (0.021) | 0.031 (0.000) | 0.030 (0.101) | 0.030 (0.075) |
|  |  | Secondary | 0.010 (0.469) | 0.010 (0.481) | 0.017 (0.336) | 0.027 (0.070) |
| Rectal | 15775/67347 | Primary | 0.002 (0.933) | 0.002 (0.936) | -0.015 (0.550) | 0.011 (0.644) |
|  |  | Secondary | -0.014 (0.472) | -0.014 (0.485) | -0.043 (0.093) | 0.004 (0.866) |
| Lung | 18082/13780 | Primary | 0.004 (0.832) | 0.004 (0.762) | -0.020 (0.477) | 0.019 (0.446) |
|  |  | Secondary | -0.006 (0.745) | -0.006 (0.750) | -0.011 (0.655) | 0.017 (0.449) |
| Pancreatic | 9254/12525 | Primary | 0.017 (0.506) | 0.017 (0.409) | 0.015 (0.702) | -0.007 (0.848) |
|  |  | Secondary | 0.021 (0.335) | 0.021 (0.264) | 0.055 (0.111) | 0.011 (0.711) |

<sup>1</sup>The primary analysis included eight SNPs (rs729876, rs1537415, rs2738058, rs12461706, rs16870060, rs2521634, rs3826782, and rs7762544). The secondary analysis included six additional SNPs (rs1122900, rs2064712, rs2070901, rs4970469, rs9982623, and rs9984417).
<sup>2</sup>The primary Mendelian randomization methods was inverse-variance weighted (IVW) MR. We used MR-PRESSO, simple median, and weighted median as secondary analyses. Betas indicate the effect estimate for the association between a one-unit increase in genetic predisposition to having chronic or aggressive periodontitis and the natural log risk for each cancer outcome.

**Figure 1.**
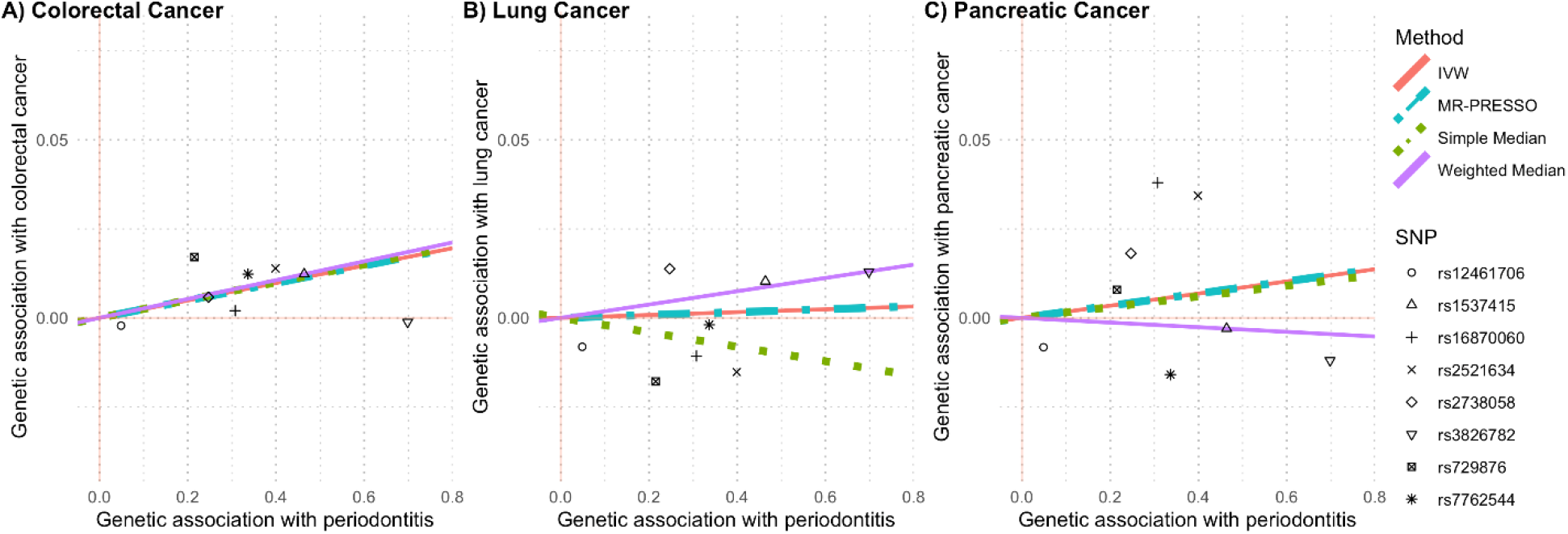
Scatterplots comparing the strength of the SNP-exposure (periodontitis) and SNP-outcome (cancer risk) associations. The lines indicate the estimated effect sizes by four Mendelian randomization methods (inverse-variance weighted (IVW), MR-PRESSO, simple median, and weighted median).

In addition to the analyses for each cancer overall, we assessed associations between genetic predisposition to having chronic or aggressive periodontitis and risk of colorectal cancer stratified by location in the large bowel (colon, rectal, distal, and proximal) and sex (Figures 2 and S1). We observed that each unit increase in the genetic predisposition index for chronic or aggressive periodontitis was associated with a 3% increased risk in colon cancer (p = 0.021), a 4% increased risk of proximal colon cancer (p = 0.013), and a 3% increased risk of colorectal cancer among females (p = 0.039; Figure 2). Each of these associations was observed with the IVW MR method and at least one alternative MR approach (MR-PRESSO for colorectal cancer, colon cancer, and proximal cancer; simple median and weighted median for colorectal cancer among females; Table S3). Additionally, whereas the primary analyses using the MR approach did not suggest significant associations with rectal cancer (p = 0.933), distal cancer (p = 0.190), or colorectal cancer in men (p = 0.174), with the MR-PRESSO method, a one-unit increase in the genetic predisposition index for chronic or aggressive periodontitis was associated with a 2% increased risk of distal colorectal cancer (p = 0.026) and a 2% increased risk of colorectal cancer in men (p = 0.010). In secondary analyses including the six additional SNPs with moderate evidence for an association with periodontitis, none of the associations assessed with any of the MR methods remained significant (Table S3).

**Figure 2.**
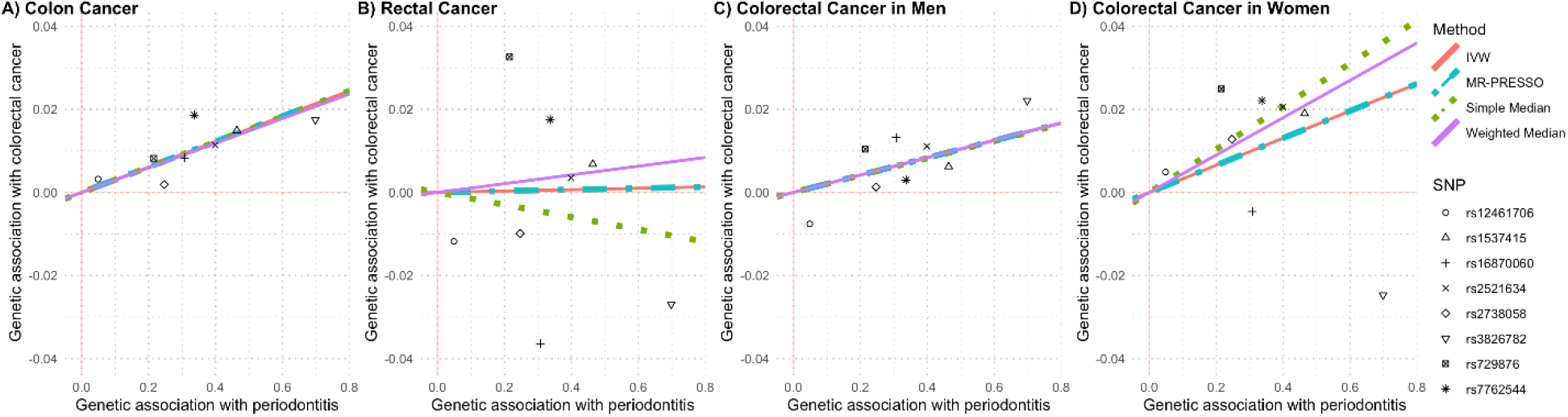
Scatterplots comparing the strength of the SNP-exposure (periodontitis) and SNP-colorectal cancer associations. The lines indicate the estimated effect sizes by four Mendelian randomization methods (inverse-variance weighted (IVW), MR-PRESSO, simple median, and weighted median).

We also investigated associations between genetic predisposition to having chronic or aggressive periodontitis and risk of lung cancer stratified by histologic type (adenocarcinoma, squamous cell, or small cell), smoker status (current or not-current), and the combination of histologic type and smoker status (Table 2). We did not observe significant associations for any of these analyses using the primary instrument with eight SNPs or with any of the MR methods (Table S4); however, in a secondary analysis including the six SNPs with moderately strong evidence for an association with periodontitis, a one-unit increase in genetic predisposition index for chronic or aggressive periodontitis was associated with a 34% decreased risk of small cell lung cancer among non-smokers (p = 0.021; Table S4). Notably, this secondary analysis included a very small number of cases (n = 64).

**Table 2.** Effect estimates for the association between genetic predisposition to having chronic or aggressive periodontitis^1^ and the risk of lung cancer by histologic sub-type and smoker status.

|  | **Overall**<br>**(N<sub>controls</sub> = 13780)** |  | **Smokers**<br>**(N<sub>controls</sub> = 9084)<sup>2</sup>** |  | **Non-smokers**<br>**(N<sub>controls</sub> = 4415)** |  |
| --- | --- | --- | --- | --- | --- | --- |
|  | N<sub>cases</sub> | B<sup>3</sup> (p-value) | N<sub>cases</sub> | β (p-value) | N<sub>cases</sub> | β (p-value) |
| Overall | 18082 | 0.004 (0.832) | 15984 | 0.002 (0.927) | 1800 | -0.021 (0.652) |
| Adenocarcinoma | 6730 | 0.025 (0.313) | 5639 | 0.029 (0.317) | 975 | 0.012 (0.839) |
| Squamous cell | 4429 | -0.010 (0.740) | 4209 | -0.019 (0.542) | 158 | -0.040 (0.773) |
| Small cell | 1853 | -0.035 (0.400) | 1761 | -0.023 (0.593) | 64 | -0.383 (0.059) |

<sup>1</sup>The inverse-variance weighted Mendelian randomization analysis included eight SNPs (rs729876, rs1537415, rs2738058, rs12461706, rs16870060, rs2521634, rs3826782, and rs7762544).
<sup>2</sup>Controls were shared across lung cancer histologic sub-types.
<sup>3</sup>Betas indicate the effect estimate for the association between a one-unit increase in genetic predisposition to having chronic or aggressive periodontitis and the natural log risk for each lung cancer outcome.

For pancreatic cancer, we observed no significant associations when we stratified by sex or smoker status (current, former, or never; Table 3). In addition, results were similar by study design (cohort or case-control; Table S5) and by dataset (i.e., PanScan I and II, PanScan III, and PanC4; data not shown). In general, the main analysis results were not substantively different than the results from the secondary analyses including the six additional SNPs (Tables S5 and S6); the only exception was for a separate analysis of the PanScan cohort studies where positive associations were observed with pancreatic cancer using both the IVW (15% increased risk; p = 0.020) and MR-PRESSO methods (15% increased risk; p = 0.012; Table S5).

**Table 3.** Effect estimates for the association between genetic predisposition to having chronic or aggressive periodontitis^1^ and the risk of pancreatic cancer by sex and smoker status using PanScan and PanC4 data.

| **Pancreatic Cancer** | **Case/Control** | **IVW**<br>**β (p value)** |
| --- | --- | --- |
| Overall | 9254/12525 | 0.017 (0.506) |
| Female | 4243/4734 | 0.036 (0.398) |
| Male | 5011/7791 | 0.003 (0.944) |
| Current smoker | 1517/1724 | 0.053 (0.438) |
| Former smoker | 3286/4982 | -0.008 (0.876) |
| Never smoker | 3314/5199 | 0.020 (0.627) |

<sup>1</sup>The inverse-variance weighted (IVW) Mendelian randomization analysis included eight SNPs (rs729876, rs1537415, rs2738058, rs12461706, rs16870060, rs2521634, rs3826782, and rs7762544).
<sup>2</sup>Betas indicate the effect estimate for the association between a one-unit increase in genetic predisposition to having chronic or aggressive periodontitis and the natural log risk for pancreatic cancer.

## Discussion

Using data from several large cancer consortia and a genetic instrument index for periodontal disease developed through a rigorous systematic selection process, we conducted the first two-sample MR assessment of periodontitis in relation to the risks of developing colorectal, lung, and pancreatic cancer. We observed evidence that a genetic predisposition to having chronic or aggressive periodontitis is associated with colorectal cancer (overall, and in a sub-analysis only including women), colon cancer, and proximal colon cancer. Conversely, our two-sample MR results were not consistent with the hypothesis that genetically predicted periodontal disease is linked to lung cancer or pancreatic cancer risk.

Our observation of an MR association between periodontitis and increased colorectal cancer risk is supported by several observational studies,^4,21,22^ though not all.^23^ Additionally, our observation that the relationship between periodontal disease and colorectal cancer risk varies by sex is supported by null associations in an all-male cohort,^24^ positive associations in one all-female cohort,^25^ and suggestive positive associations in another small all-female cohort (n = 19 participants with colorectal cancer);^26^ however, a large cohort study with clinical measurements for periodontal disease reported similar positive associations in men and women.^27^ More studies will need to examine the role of sex in the association between periodontal disease and colorectal cancer.

Plausible causal mechanisms linking periodontal disease to colorectal cancer incidence may involve inflammatory processes or oral microbiome shifts (dysbiosis) that migrate to extra oral sites.^28,29^ For example, the gram-negative *Fusobacteria* is among the quantitatively dominant microorganisms in dental plaque;^30^ it interacts with inflammatory processes associated with periodontal disease; and it has been identified in colorectal cancer tissue.^30–32^ Notably, the proportion of colorectal cancer cases with high *Fusobacteria* varies by location (generally observed more in proximal versus distal cases).^33–35^ Furthermore, microbiota organization (e.g., presence of a bacterial biofilm) is particularly associated with proximal colon cancer compared to distal colon cancer.^36^ These observations, along with previous studies indicating that multiple environmental factors and mutation profiles have differential associations by cancer location in the large bowel,^37–40^ support our finding that genetic predisposition to having chronic or aggressive periodontal disease may be more likely to influence proximal colon cancer risk than distal colon cancer or rectal cancer risk.

In contrast to meta-analyses of observational studies suggesting that periodontal disease is associated with the risk of lung and pancreatic cancer,^6–8,41,42^ our primary analyses did not indicate that there were significant associations between a genetic predisposition to having chronic or aggressive periodontal disease and the risk of either lung or pancreatic cancer. Given that the effect estimates for colorectal cancer and pancreatic cancer were similar but we had more colorectal cancer cases, it is possible that our pancreatic cancer analyses were underpowered. Additionally, residual confounding by smoking status could explain some of the differences in our results and results from certain observational studies (especially due to the null results in our secondary analyses including only non-smokers).^6^ The significant association we observed in one of our analyses with the secondary genetic instrument using only data from PanScan cohort study participants could be a chance finding or it could suggest that other pathways are involved that we failed to capture with our existing primary instrument. Based on our overall results, it is possible that periodontal disease is not causally involved with lung or pancreatic cancer initiation and may instead be linked with cancer progression. This hypothesis would be supported by evidence that cancer progression is related to increased presence of certain oral bacteria common among individuals with periodontal disease^43^ or is affected by oxidative stress, inflammatory, or immunological responses associated with periodontal disease.^44–46^ More research using markers of periodontal disease that may affect cancer progression could provide more insight into this alternative scenario.

As with all MR studies, one limitation of our analysis is the potential for violations of the MR assumptions. For example, we could not directly test for the presence (or impact of) directional pleiotropy using MR-Egger due to the small number of genetic variants in our instrument. However, the overall consistency of our primary analyses using the IVW method and secondary analyses using different MR methods (MR-PRESSO, simple median, and weighted median – an approach that is less biased by the presence of directional pleiotropy^47^) suggests both that directional pleiotropy is unlikely to completely explain the results and that outliers were unlikely to substantially affect the results. Bias could also arise due to assortative mating (such that the parental genotypes are correlated).^48,49^ Additionally, the primary phenotypes associated with at least several of the SNPs used in the analysis (e.g., rs2738058, rs12461706, and rs2070901) involve inflammatory pathways. Since inflammation processes are likely on a putative causal pathway between periodontal disease and cancer risk, our choice of SNPs may introduce vertical pleiotropy and potentially strengthen the genetic instrument. Finally, we may have observed significant associations by chance due to the multiple comparisons made, we did not have data stratified by smoker status for colorectal data or smoking data that distinguished former versus never smokers for lung cancer, and our study results are only generalizable to individuals of European ancestry.

Strengths of our study include the large number of cancer cases included in each analysis, our MR approach that limits the potential for confounding or reverse causation, and our systematic approach to SNP selection for inclusion in our genetic instrument. Previous two-sample MR analyses that used genetic instruments for periodontal disease (examining non-cancer outcomes) included SNPs identified from single GWAS articles without clear justification for using those specific articles and SNPs.^15,17^ Another two-sample MR analysis included SNPs that were not significantly associated with periodontitis as well as SNPs that were significantly associated with the autoimmune outcomes (rheumatoid arthritis and systemic lupus erythematosus) in GWAS (potentially introducing bias due to violations of the MR assumptions).^16,50^ In contrast, we examined the strength of the evidence for an association of each SNP with periodontal disease based on objective criteria (e.g., inclusion of validation and replication cohorts, definition of periodontal disease). We also used secondary analyses and sensitivity analyses to quantitatively assess our assumptions about these criteria. Finally, our inclusion of SNPs associated with aggressive (early onset) periodontitis may reflect the risk of periodontal disease only, rather than possible shared risk factors of periodontal disease and cancer.

Our two-sample MR analysis utilizing a systematically developed genetic instrument suggests that a genetic predisposition to having chronic or aggressive periodontal disease may be associated with colorectal cancer risk. Additionally, our results suggest confounding is unlikely to fully explain previous observational studies’ claims for an association between periodontal disease and colorectal cancer. Our results were not consistent with the hypothesis that a genetic predisposition to having periodontal disease is associated with lung or pancreatic cancer risk; however, we cannot entirely rule out the possibility that periodontal disease is associated with either of these cancers. Taken together, our results suggest that increased attention to preventative oral health measures and increased cancer surveillance of patients with periodontitis may be warranted. Future research is needed to further elucidate biological pathways underlying the associations between periodontitis and cancer risk.

## Methods

### Genetic instrument for periodontal disease

We determined two genetic instruments for periodontal disease based on a systematic evaluation of the strength of the GWAS evidence for associations between individual SNPs and chronic, aggressive, and/or severe periodontal disease (Table S1).^51–58^ Aggressive periodontitis was defined by percentage bone loss affecting multiple teeth in adults <36 years of age as determined by full-mouth dental radiographs.^51–53,59^ Chronic and severe periodontitis were defined by measures such as age- and sex-specific groups of attachment loss ≥ 4⍰mm;^51,52^ self-reported gum surgery;^55^ or probing depth, clinical attachment level, plaque index, gingival index, and bleeding for multiple teeth.^54,55,60^

There were eight SNPs in our primary instrument, of which five had very strong evidence for an association with periodontal disease and three had strong evidence. We considered the evidence very strong if the association with periodontitis met the genome-wide significance threshold of p < 5 × 10^−8^ in a pooled analysis of multiple cohorts (focusing on populations of European descent to match the population demographics of our outcome data). Six SNPs (rs729876, rs1537415, rs2738058, rs2978951, rs4284742, and rs16870060) met this definition.^51–53^ Four of these SNPs (rs729876, rs2738058, rs4284742, and rs16870060) were used as the instrumental variables for periodontitis in a previous MR study on hypertension; the authors did not include rs1537415 because it is palindromic and did not have a proxy SNP in Caucasians.^17^ We included the palindromic rs1537415 in our primary analysis because the minor allele frequency was 0.42 and we considered this sufficiently different from 0.50; however, we also conducted sensitivity analyses excluding this allele. Since rs2738058 and rs2978951 were in linkage disequilibrium (r^2^ = 0.27; p < 0.001 for European populations),^61^ and since rs2738058 had a larger estimated effect estimate and smaller p-value (odds ratio = 1.28 versus 1.25; p = 6.78 × 10^−10^ versus 2.06 × 10^−8^, respectively), we excluded rs2978951.^51^ Similarly, we included rs12461706 instead of rs4284742 because the two SNPs were in linkage disequilibrium (r^2^ = 0.19; p < 0.001 for European populations),^61^ one study observed that rs12461706 was significantly associated with periodontitis and loose teeth (p = 3.9 × 10^−9^ in a combined analysis; consistent effect estimates observed in two study samples),^55^ the p-values reported in PhenoScanner for associations with dental health traits were smaller for rs12461706 than for rs4284742,^62,62^ and substituting rs12461706 substantially increased the *I*^2^_*GX*_ value (0.62 versus 0.93). In addition to the five SNPs with very strong evidence for an association, we identified three SNPs (rs2521634, rs3826782, and rs7762544) with strong evidence for an association. These SNPs were positively associated with chronic periodontitis in one cohort (p < 5×10^−6^), nominally positively associated (p < 0.05) with severe chronic periodontitis in an independent replication cohort, and positively associated (p < 5 × 10^−6^) with chronic periodontitis in a meta-analysis of over 5000 European American individuals in these cohorts.^54^

As a secondary analysis, we identified seven SNPs (rs11084095, rs1122900, rs2064712, rs2070901, rs4970469, rs9982623, and rs9984417) with moderate evidence for an association with periodontitis (associated with periodontitis with a threshold of p < 5 × 10^−6^ in a pooled analysis of multiple cohorts but not associated with periodontitis in any single cohort with a threshold of p < 5 × 10^−6^).^51,52^ Three of these SNPs (rs1122900, rs2070901, and rs4970469) were associated with aggressive periodontitis (p < 5 × 10^−6^) in the combined results from a discovery meta-analysis and replication cohort.^51^ The other four SNPs (rs11084095, rs2064712, rs9982623, and rs9984417) were associated with aggressive and chronic periodontitis (p < 5 × 10^−6^) in pooled cohorts.^52^ One of these SNPs (rs11084095) was in linkage disequilibrium with a SNP we identified as having very strong evidence for an association (rs12461706) so we excluded rs11084095 (r^2^ = 0.99; p < 0.001 for European populations).^61^ Our secondary instrument included 14 SNPs (eight used in the primary instrument, plus these six; Table S1).

We assessed whether any of our eight primary SNPs (those with very strong or strong evidence for an association with periodontal disease) or six secondary SNPs (those with moderate evidence for an association with periodontal disease) were potentially pleiotropic, in mutual linkage disequilibrium, accounted for population stratification, or were problematic to harmonize. Of the 14 SNPs, only rs2070901, rs2738058, and rs12461706 were associated (p < 5 × 10^−8^) with any diseases or traits other than periodontitis or other oral health measures.^63^ Notably, the associated traits were primarily related to inflammatory processes (e.g., white blood cell differential) that may be part of the putative causal pathways between periodontal disease and cancer risk (thus introducing vertical pleiotropy and potentially strengthening the genetic instrument). None of the included SNPs was in linkage disequilibrium with any other included SNP.^61^ Similarly, population stratification seems unlikely to be a major confounder of the GWAS results; the participants included in the analyses for all of the SNPs are primarily from populations of European descent^51–54^ and most of the studies explicitly considered population stratification.^51,54,55^ All genetic variants were positively associated with periodontitis. We verified that the genetic variant was the same in each dataset for palindromic alleles (rs1537415, rs9984417, and rs12461706), and we conducted sensitivity analyses removing each of these SNPs (one at a time) to ensure that the effect estimates were not driven by issues related to harmonization.

### Summary-level data for lung, colorectal, and pancreatic cancer

We used summary-level genetic data for colorectal cancer (overall, by location in the large bowel, and by sex) from 125,478 participants (including nearly 58,131 colorectal cancer and advanced adenoma patients) in the Genetics and Epidemiology of Colorectal Cancer Consortium (GECCO; 13 studies), Colon Cancer Family Registry (CCFR), and Colorectal Transdisciplinary study (CORECT);^64–66^ lung cancer (overall, by histologic type, and by smoker status [current/non-current]) from 31,862 participants (including 18,082 patients) in the International Lung Cancer Consortium (ILCCO; 26 studies included);^67,68^ pancreatic cancer (overall, by sex, smoker status, data source [pancreatic cancer consortium], and study design) for 13,823 participants (including 5090 patients) in PanScan I and II (12 cohort and eight case-control studies) and PanScan III (15 cohorts, two case series, and one case-control study);^65,69^ and pancreatic cancer (overall, by sex, and by smoker status) in 7956 participants in the Pancreatic Cancer Case-Control Consortium (PanC4; including 4164 patients).^70–72^ Choices for secondary analyses (e.g., not assessing colorectal cancer associations by smoker status) were due to data availability. All cancer data for our analysis came from individuals of European ancestry. All studies participating in each consortium obtained informed consent from participants and approval from the relevant ethical review boards. None of the study samples that contributed genetic data for lung, colorectal, or pancreatic cancer overlapped with study samples that contributed data for the periodontitis GWAS.

Genotyping and imputation methods for each consortium have been described previously. Briefly, for GECCO, genotyping was performed with one of the Illumina 1536 GoldenGate assay (Illumina, Inc, San Diego, CA), the Affymetrix GeneChip Human Mapping 100K and 500K Array Set (Affymetrix, Inc, Santa Clara, CA), or a 10K nonsynonymous SNP chip.^64^ SNPs were excluded from genotyping if the call rate was <98%, there was a low minor allele frequency (<1%), or there was a lack of Hardy-Weinberg equilibrium in controls (p < 10^−4^). Imputed data were included for autosomal SNPs with minor allele frequency ≥1% and high imputation accuracy (R^2^ > 0.3). The reference population included Utah residents of Northern and Western European ancestry from HapMap II.^73^ For ILCCO, quality control procedures for genotyping with the OncoArray included steps to account for duplicates, related individuals, Hardy-Weinberg equilibrium, and call rates.^74^ Imputation was conducted with the 1000 Genomes Project Phase 3 as a reference.^75^ For PanScan I, II, and III, genotyping was performed at the National Cancer Institute Cancer Genomics Research Laboratory with Illumina HumanHap series arrays (Illumina HumanHap550 Infinium II, Human 610-Quad) for PanScan I and II, respectively, and the Illumina Omni series arrays (OmniExpress, Omni1M, Omni2.5 and Omni5M) for PanScan III. Quality control procedures accounted for duplicates, related individuals, population admixture, Hardy-Weinberg equilibrium, and call rates. PanC4 was genotyped at the Center for Inherited Disease Research.^76^ Genotyping was performed with the Illumina HumanOmniExpressExome-8v1 array.^72^ Imputation for both PanScan and PanC4 was conducted using the 1000 Genomes Phase 3, Release 1 reference data set and IMPUTE2.^77–80^ Imputation analyses were adjusted for age in decade, sex, and top eigenvectors (five for PanScan; nine for PanC4) from principal components analysis to control for ancestry. PanScan analyses were also adjusted for study and geographic region of the parent studies.

### Statistical analyses

We first estimated the association of genetic predisposition index for having chronic or aggressive periodontal disease on colorectal, lung, and pancreatic cancer risk using the random-effects inverse-variance weighted (IVW) method. The IVW method is commonly used because it has greater power to detect associations than many other MR methods and because the results are equivalent to the two-stage least squares method that would be obtained using individual-level data. Since the IVW method assumes that each genetic variant is a valid instrumental variable and it forces a zero intercept in the regression slope, the IVW method assumes either no pleiotropy or balanced directional pleiotropy and that any balanced pleiotropic effects of a genetic variant are independent of the association with the exposure.^47,81,82^ We also considered three other MR estimation methods with different assumptions to evaluate the robustness of the IVW findings. The first alternative MR method we considered was MR-PRESSO. MR-PRESSO identifies and excludes outlier SNPs using the residual sum of squares heterogeneity measure.^83^ Although this method is efficient if each SNP is a valid instrument and it can help handle directional pleiotropy, it is prone to false positives if multiple SNPs are invalid.^81^ The other two MR methods we considered (simple median and weighted median) assume that the majority of SNPs are valid.^47,81,84^ We did not use MR-Egger (a common MR method that examines directional pleiotropy) because the consistency of MR-Egger effect estimates increases as the number of genetic variants increases.^85^ Specifically, the MR-Egger regression models would not have sufficient power to detect true associations (with 15 SNPs, MR-Egger has power = 0.25 versus the IVW method that has power = 0.78).^85^

For each MR method, our primary analyses included only the eight SNPs with very strong or strong evidence for an association with periodontal disease. We also ran secondary analyses adding in the six SNPs with moderate evidence for an association with periodontal disease. We quantitatively assessed violations of the ‘NO Measurement Error’ (NOME) assumption using the *I*^2^_*GX*_ statistic (where values between 0.9 and 1 suggest that violations are negligible and that the uncertainty in the SNP-exposure associations are substantially smaller than the underlying heterogeneity in these associations).^86,87^ We assessed heterogeneity using the *I*^2^ statistic (where values >50% indicate potential horizontal pleiotropy). All statistical analyses were performed in R (version 3.6.2) with packages *MendelianRandomization* (version 0.4.2), *MRPRESSO* (version 1.0). We constructed plots showing the genetic associations of the SNPs with periodontitis (natural log values of the odds ratios shown in Table S1) versus the genetic associations of the SNPs with cancer (natural log values of the odds ratios provided by each consortium) using *ggplot2* (slope represents the beta values for the MR models). Default parameters were used for each analysis. Since we only used de-identified data, we did not need Institutional Review Board approval for our analysis.

## Data Availability

The data for the main analyses conducted in this article are available through dbGAP or by submitting a request to each of the consortia.

https://www.ncbi.nlm.nih.gov/projects/gap/cgi-bin/analysis.cgi?study_id=phs001273.v3.p2&phv=282571&phd=7215&pha=4930&pht=6171&phvf=&phdf=&phaf=&phtf=&dssp=1&consent=&temp=1

https://www.ncbi.nlm.nih.gov/projects/gap/cgi-bin/study.cgi?study_id=phs000648.v1.p1

https://www.ncbi.nlm.nih.gov/projects/gap/cgi-bin/study.cgi?study_id=phs001499.v1.p1

https://www.ncbi.nlm.nih.gov/projects/gap/cgi-bin/study.cgi?study_id=phs001078.v1.p1

## Funding

This work was supported the AACR-Johnson & Johnson Lung Cancer Innovation Science Grant Number 18-90-52-MICH. LC was supported by the National Institute of Child Health & Human Development at the National Institutes of Health (grant number K12HD092535). KKT was supported by Cancer Research UK (grant number: C18281/A29019). CA was a Research Scholar supported by Cancer Prevention Research Institute of Texas grant RR170048. The supplement lists funding for the cancer consortia that provided genetic data for this analysis. None of the funders had any role in this analysis or interpretation of the data; the writing of the manuscript; or the decision to submit this manuscript for publication.

## Acknowledgements

The authors would like to thank the study participants and staff of the Seattle Colon Cancer Family Registry and the Hormones and Colon Cancer study (CORE Studies). They would also like to thank Kimon Divaris for helpful guidance on selecting SNPs and genes that are considered causal in periodontal disease.

## Author contributions

LC wrote the first draft of the manuscript. MR conducted the primary analysis. KKT and EB helped with the analysis. DSM conceived of the analysis, acquired the data from the various consortia, and oversaw all aspects of the work. All authors contributed to the data interpretation and manuscript revisions.

## Author disclosures

No authors have any conflicts of interest to disclose.

## Data availability statement

Data underlying this article are available through dbGAP:

https://www.ncbi.nlm.nih.gov/projects/gap/cgi-bin/analysis.cgi?study_id=phs001273.v3.p2&phv=282571&phd=7215&pha=4930&pht=6171&phvf=&phdf=&phaf=&phtf=&dssp=1&consent=&temp=1; https://www.ncbi.nlm.nih.gov/projects/gap/cgi-bin/study.cgi?study_id=phs000206.v3.p2; https://www.ncbi.nlm.nih.gov/projects/gap/cgi-bin/study.cgi?study_id=phs000648.v1.p1; https://www.ncbi.nlm.nih.gov/projects/gap/cgi-bin/study.cgi?study_id=phs001078.v1.p1; and https://www.ncbi.nlm.nih.gov/projects/gap/cgi-bin/study.cgi?study_id=phs001499.v1.p1. Data unavailable in dbGaP can be requested from the respective consortia.

## Notes

### Competing Interest Statement

The authors have declared no competing interest.

### Funding Statement

This work was supported the AACR-Johnson & Johnson Lung Cancer Innovation Science Grant Number 18-90-52-MICH. Laura Corlin was supported by the National Institute of Child Health & Human Development at the National Institutes of Health (grant number K12HD092535). Konstantinos K Tsilidis is supported by Cancer Research UK (grant number: C18281/A29019). Christopher Amos is a Research Scholar supported by Cancer Prevention Research Institute of Texas grant RR170048.

### Author Declarations

Consortium nameEthics Committee / Institutional Review Board (IRB) Decision made by ethical oversight body PanScan studiesNCI Special Studies IRB (SSIRB)Approved under secondary aims PanC4 IRB of Johns Hopkins MedicineWaived (de-identified data) ILCCOIRB of Baylor College of MedicineWaived (de-identified data) GECCOIRB of Fred Hutchinson Cancer Research CenterApproved CCFRIRB of Fred Hutchinson Cancer Research CenterApproved CCFRMount Sinai Hospital Research Ethics BoardApproved CCFRUniversity of Hawaii Human Studies ProgramApproved CCFRUniversity of Melbourne Health Sciences Human Ethics Sub-CommitteeApproved CCFRMayo Clinic IRBApproved CCFRCSMC Institutional Review BoardsApproved CORECTIRB of University of Southern California Health SciencesApproved

