## Supplemental Materials for "Two-sample Mendelian randomization analysis of associations between periodontal disease and risk of colorectal, lung, and pancreatic cancers"

**Supplemental Figures and Tables**

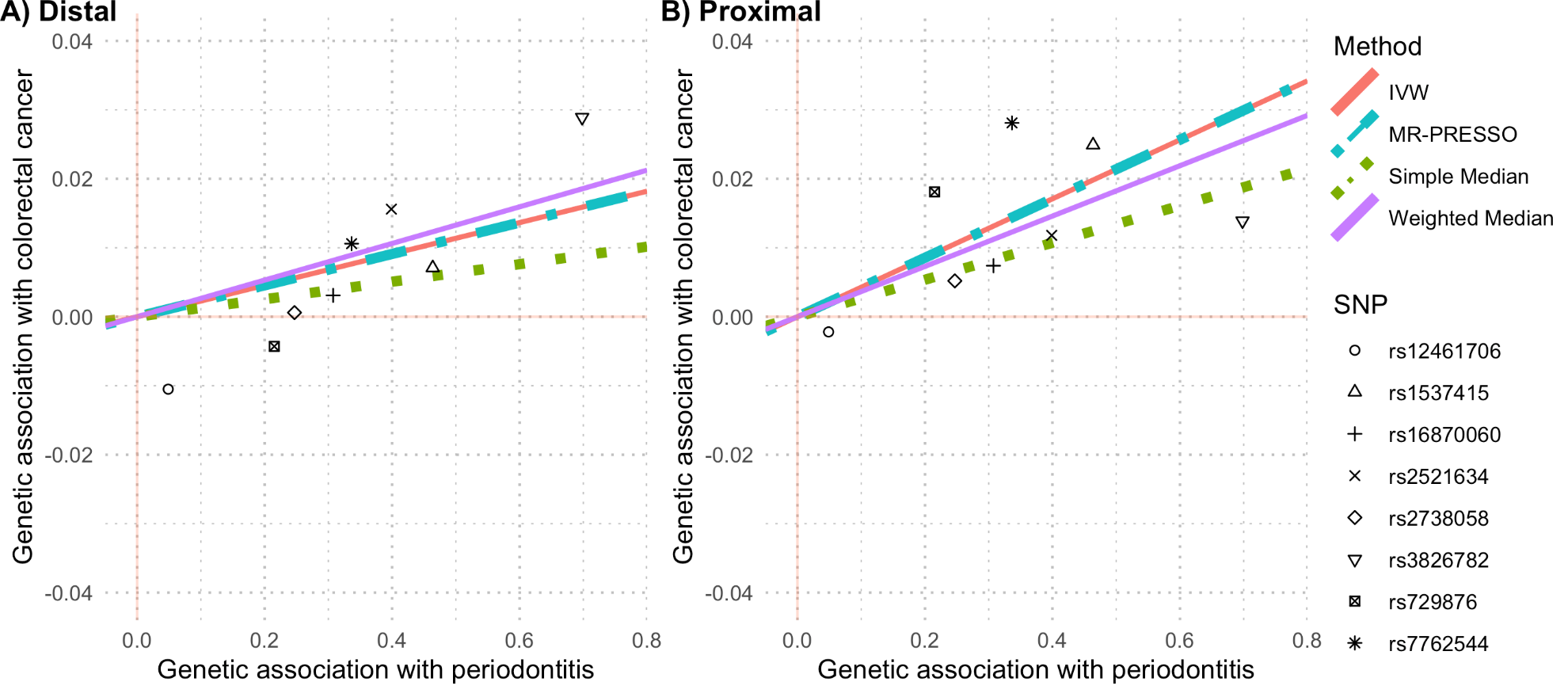

**Figure S1.** Scatterplots comparing the strength of the SNP-exposure (periodontitis) and SNP-colorectal cancer associations. The lines indicate the estimated effect sizes by four Mendelian randomization methods (inverse‐variance weighted (IVW), MR-PRESSO, simple median, and weighted median).

**Table S1.** Single nucleotide polymorphisms (SNPs) used as instrumental variables based on their association with periodontal disease

| Evidence | rsID | Chromosome | Nearest gene (or pseudogene) | Effect allele | Non-effect allele | OR (95% CI) | Effect allele frequency (periodontal disease) | Effect allele frequency (colorectal cancer, lung cancer, and pancreatic cancer) | Periodontal disease definition^5^ and reference number |
| --- | --- | --- | --- | --- | --- | --- | --- | --- | --- |
| Very strong^1^ | rs729876 | 16 | *SHISA9* | T | C | 1.24 (1.15-1.34) | 0.79-0.84 | 0.80-0.81 | Aggressive or chronic (1) |
|  | rs1537415 | 9 | *GLT6D1* | G | C | 1.59 (1.36-1.86) | 0.50-0.62 | 0.59-0.60 | Aggressive (2) |
|  | rs2738058 | 8 | *DEFA1A3* | T | C | 1.28 (1.18-1.38) | 0.43 | 0.42-0.44 | Aggressive or chronic (3) |
|  | rs12461706 | 19 | *SIGLEC5* | T | A | 1.05 (1.03, 1.07)^4^ | 0.40 | 0.39-0.41 | Chronic (5) |
|  | rs16870060 | 8 | *MTND1P5* | G | T | 1.36 (1.23-1.51) | 0.89-0.94 | 0.91 | Aggressive or chronic (1) |
| Strong^2^ | rs2521634 | 7 | *NPY* | A | G | 1.49 (1.28-1.73) | 0.20-0.26 | 0.24 | Chronic (6) |
|  | rs3826782 | 19 | *EMR1* | A | G | 2.01 (1.52-2.65) | 0.04-0.05 | 0.09-0.11 | Chronic (6) |
|  | rs7762544 | 6 | *NCR2* | G | A | 1.40 (1.24-1.59) | 0.16-0.21 | 0.19-0.20 | Chronic (6) |
| Moderate^3^ | rs1122900 | 5 | *CTD-2353F22.1* | A | C | 1.27 (1.16-1.40) | 0.40 | 0.42 | Aggressive or chronic (3) |
|  | rs2064712 | 6 | *AL109933.3-AL391361.2* | A | G | 1.24 (1.14-1.35) | 0.09-0.19 | 0.11-0.16 | Aggressive or chronic (1) |
|  | rs2070901 | 1 | *FCER1G* | T | G | 1.29 (1.16-1.44) | 0.24 | 0.26-0.27 | Aggressive or chronic (3) |
|  | rs4970469 | 1 | *OSTCP2* | G | A | 1.52 (1.29-1.81) | 0.90 | 0.89-0.90 | Aggressive or chronic (3) |
|  | rs9982623 | 21 | *MCM3AP* | C | T | 1.24 (1.14-1.35) | 0.86-0.89 | 0.86-0.88 | Aggressive or chronic (1) |
|  | rs9984417 | 21 | *MAPK6PS2-AP000959.2* | T | A | 1.15 (1.09-1.23) | 0.61-0.68 | 0.60-0.61 | Aggressive or chronic (1) |

^1^Very strong evidence: the association with periodontitis met the genome-wide significance threshold of p < 5 x 10^-8^ in a pooled analysis of multiple cohorts (for each level of evidence, focusing on populations of European descent to match the population demographics of our outcome data)
^2^Strong evidence: associated with periodontitis in at least one cohort with a threshold of p < 5 x 10^-6^, same direction of association in an independent cohort with a threshold of p < 0.05, same direction of association in a meta-analysis of these cohorts with a threshold of p < 5 x 10^-6^
^3^Moderate evidence: associated with periodontitis with a threshold of p < 5 x 10^-6^ in a pooled analysis of multiple cohorts but not associated with periodontitis in any single cohort with a threshold of p < 5 x 10^-6
4^The 95% confidence interval for this SNP was calculated using the p-value and sample size reported in the source article.
^5^Aggressive periodontitis was defined by percentage bone loss affecting multiple teeth in adults <36 years of age as determined by full-mouth dental radiographs.^1–4^ Chronic and severe periodontitis were defined by measures such as age- and sex-specific groups of attachment loss ≥ 4 mm;^1,3^ self-reported gum surgery;^5^ or probing depth, clinical attachment level, plaque index, gingival index, and bleeding for multiple teeth.^5–7^

**Table S2**. Effect estimates for the association between genetic predisposition to having chronic or aggressive periodontitis and the risk of colorectal, lung, and pancreatic cancer by MR approach and genetic instrument sensitivity analysis.

| Outcome | Instrument^1^ | IVW^2^  β (p-value) | MR-PRESSO β (p-value) | Simple median β (p-value) | Weighted median β (p-value) |
| --- | --- | --- | --- | --- | --- |
| Colorectal (N_cases_= 58131, N_controls =_ 67347) | Primary | 0.025 (0.026) | 0.025 (0.010) | 0.025 (0.115) | 0.027 (0.063) |
|  | Primary excluding rs1537415 | 0.024 (0.077) | 0.024 (0.041) | 0.023 (0.246) | 0.027 (0.114) |
|  | Primary excluding rs3826782 | 0.031 (0.012) | 0.031 (0.002) | 0.027 (0.137) | 0.030 (0.052) |
|  | Primary excluding rs12461706 | 0.025 (0.025) | 0.025 (0.015) | 0.027 (0.090) | 0.027 (0.063) |
|  | Primary excluding rs1537415 and rs12461706 | 0.024 (0.074) | 0.024 (0.058) | 0.029 (0.094) | 0.027 (0.105) |
|  | Secondary | 0.006 (0.699) | 0.016 (0.106) | 0.002 (0.875) | 0.025 (0.054) |
|  | Secondary excluding rs9984417 | 0.006 (0.692) | 0.017 (0.111) | 0.007 (0.682) | 0.025 (0.057) |
| Lung  (N_cases_ = 18082,  N_controls_ = 13780) | Primary | 0.004 (0.832) | 0.004 (0.762) | -0.020 (0.477) | 0.019 (0.446) |
|  | Primary excluding rs1537415 | -0.003 (0.878) | -0.003 (0.829) | -0.035 (0.320) | 0.001 (0.966) |
|  | Primary excluding rs3826782 | -0.001 (0.950) | -0.001 (0.932) | -0.035 (0.330) | 0.003 (0.905) |
|  | Primary excluding rs12461706 | 0.005 (0.809) | 0.005 (0.742) | -0.006 (0.833) | 0.019 (0.440) |
|  | Primary excluding rs1537415 and rs12461706 | -0.003 (0.904) | -0.003 (0.873) | -0.020 (0.505) | 0.001 (0.961) |
|  | Secondary | -0.006 (0.745) | -0.006 (0.750) | -0.011 (0.655) | 0.017 (0.449) |
|  | Secondary excluding rs9984417 | -0.007 (0.704) | -0.007 (0.711) | -0.017 (0.532) | 0.015 (0.496) |
| Pancreatic (PanScan)  (N_cases_ = 5090,  N_controls_ = 8733) | Primary | 0.031 (0.372) | 0.031 (0.189) | 0.031 (0.537) | 0.026 (0.550) |
|  | Primary excluding rs1537415 | 0.032 (0.428) | 0.032 (0.276) | 0.035 (0.583) | 0.031 (0.561) |
|  | Primary excluding rs3826782 | 0.037 (0.344) | 0.037 (0.195) | 0.035 (0.574) | 0.033 (0.516) |
|  | Primary excluding rs12461706 | 0.031 (0.364) | 0.031 (0.215) | 0.035 (0.471) | 0.026 (0.545) |
|  | Primary excluding rs1537415 and rs12461706 | 0.033 (0.418) | 0.033 (0.309) | 0.044 (0.416) | 0.031 (0.550) |
|  | Secondary | 0.033 (0.262) | 0.033 (0.170) | 0.044 (0.312) | 0.026 (0.513) |
|  | Secondary excluding rs9984417 | 0.033 (0.262) | 0.033 (0.170) | 0.044 (0.312) | 0.026 (0.513) |
| Pancreatic (PanC4)  (N_cases_ = 4164,  N_controls_ = 3792) | Primary | 0.000 (0.994) | 0.000 (0.992) | -0.020 (0.738) | -0.030 (0.548) |
|  | Primary excluding rs1537415 | 0.020 (0.665) | 0.020 (0.568) | 0.008 (0.913) | 0.012 (0.843) |
|  | Primary excluding rs3826782 | 0.017 (0.708) | 0.017 (0.620) | 0.008 (0.912) | 0.002 (0.970) |
|  | Primary excluding rs12461706 | 0.000 (0.991) | 0.000 (0.989) | 0.008 (0.893) | -0.029 (0.550) |
|  | Primary excluding rs1537415 and rs12461706 | 0.021 (0.648) | 0.021 (0.577) | 0.031 (0.613) | 0.012 (0.834) |
|  | Secondary | 0.006 (0.855) | 0.006 (0.785) | 0.030 (0.551) | -0.042 (0.353) |
|  | Secondary excluding rs9984417 | 0.006 (0.855) | 0.006 (0.785) | 0.030 (0.551) | -0.042 (0.353) |

^1^The primary analysis included eight SNPs (rs729876, rs1537415, rs2738058, rs12461706, rs16870060, rs2521634, rs3826782, and rs7762544). The secondary analysis included six additional SNPs (rs1122900, rs2064712, rs2070901, rs4970469, rs9982623, and rs9984417).
^2^The primary Mendelian randomization methods was inverse‐variance weighted (IVW) MR. We used MR-PRESSO, simple median, and weighted median as secondary analyses. Betas indicate the effect estimate for the association between a one-unit increase in genetic predisposition to having chronic or aggressive periodontitis and the natural log risk for each outcome.

**Table S3.** Effect estimates for the association between genetic predisposition to having chronic or aggressive periodontitis and the risk of colorectal cancer by histologic type, sex, genetic instrument, and MR approach.

| Outcome | N_cases_/N_controls_ | Instrument^1^ | IVW^2^  β (p-value) | MR-PRESSO β (p-value) | Simple median β (p-value) | Weighted median β (p-value) |
| --- | --- | --- | --- | --- | --- | --- |
| All colorectal cancer | 58131/67347 | Primary | 0.025 (0.026) | 0.025 (0.010) | 0.025 (0.115) | 0.027 (0.063) |
|  |  | Secondary | 0.006 (0.699) | 0.016 (0.106) | 0.002 (0.875) | 0.025 (0.054) |
| Colon cancer | 31083/67347 | Primary | 0.031 (0.021) | 0.031 (0.000) | 0.030 (0.101) | 0.030 (0.075) |
|  |  | Secondary | 0.010 (0.469) | 0.010 (0.481) | 0.017 (0.336) | 0.027 (0.070) |
| Rectal cancer | 15775/67347 | Primary | 0.002 (0.933) | 0.002 (0.936) | -0.015 (0.550) | 0.011 (0.644) |
|  |  | Secondary | -0.014 (0.472) | -0.014 (0.485) | -0.043 (0.093) | 0.004 (0.866) |
| Distal cancer | 15306/67347 | Primary | 0.023 (0.190) | 0.023 (0.026) | 0.013 (0.593) | 0.027 (0.215) |
|  |  | Secondary | -0.001 (0.972) | -0.001 (0.973) | -0.009 (0.707) | 0.014 (0.471) |
| Proximal cancer | 13857/67347 | Primary | 0.043 (0.013) | 0.043 (0.002) | 0.027 (0.279) | 0.036 (0.100) |
|  |  | Secondary | 0.020 (0.212) | 0.020 (0.234) | 0.020 (0.360) | 0.029 (0.139) |
| Colorectal cancer in females | 26843/32820 | Primary | 0.033 (0.039) | 0.033 (0.065) | 0.052 (0.033) | 0.045 (0.037) |
|  |  | Secondary | 0.016 (0.333) | 0.016 (0.351) | 0.012 (0.574) | 0.031 (0.104) |
| Colorectal cancer in males | 31288/34527 | Primary | 0.021 (0.174) | 0.021 (0.010) | 0.021 (0.320) | 0.021 (0.266) |
|  |  | Secondary | -0.002 (0.923) | -0.002 (0.924) | 0.007 (0.731) | 0.016 (0.384) |

^1^The primary analysis included eight SNPs (rs729876, rs1537415, rs2738058, rs12461706, rs16870060, rs2521634, rs3826782, and rs7762544). The secondary analysis included six additional SNPs (rs1122900, rs2064712, rs2070901, rs4970469, rs9982623, and rs9984417).
^2^The primary Mendelian randomization methods was inverse‐variance weighted (IVW) MR. We used MR-PRESSO, simple median, and weighted median as secondary analyses. Betas indicate the effect estimate for the association between a one-unit increase in genetic predisposition to having chronic or aggressive periodontitis and the natural log risk for each outcome.

**Table S4**. Effect estimates for the association between genetic predisposition to having chronic or aggressive periodontitis and the risk of lung cancer by cancer location, smoker status, genetic instrument, and MR approach.

| Outcome | N_cases_/N_controls_ | Instrument^1^ | IVW^2^  β (p-value) | MR-PRESSO β (p-value) | Simple median β (p-value) | Weighted median β (p-value) |
| --- | --- | --- | --- | --- | --- | --- |
| Adenocarcinoma | 6730/ 13780 | Primary | 0.025 (0.313) | 0.025 (0.248) | 0.035 (0.320) | 0.036 (0.266) |
|  |  | Secondary | 0.012 (0.708) | 0.031 (0.222) | 0.040 (0.201) | 0.038 (0.196) |
| Squamous cell | 4429/ 13780 | Primary | -0.010 (0.740) | -0.010 (0.706) | -0.028 (0.498) | 0.011 (0.769) |
|  |  | Secondary | -0.019 (0.446) | -0.019 (0.449) | -0.028 (0.454) | 0.008 (0.807) |
| Small cell | 1853/ 13780 | Primary | -0.035 (0.400) | -0.035 (0.205) | -0.033 (0.602) | -0.033 (0.545) |
|  |  | Secondary | -0.019 (0.589) | -0.019 (0.416) | -0.032 (0.549) | -0.034 (0.486) |
| Smokers | 15984/9084 | Primary | 0.002 (0.927) | 0.002 (0.924) | 0.011 (0.758) | 0.020 (0.506) |
|  |  | Secondary | 0.001 (0.972) | 0.001 (0.970) | 0.011 (0.715) | 0.018 (0.519) |
| Non-smokers | 1800/ 4415 | Primary | -0.021 (0.652) | -0.021 (0.615) | -0.051 (0.511) | 0.010 (0.885) |
|  |  | Secondary | -0.042 (0.394) | -0.042 (0.409) | -0.022 (0.746) | 0.006 (0.915) |
| Smokers with adenocarcinoma | 5639/ 9084 | Primary | 0.029 (0.317) | 0.029 (0.350) | 0.034 (0.418) | 0.042 (0.274) |
|  |  | Secondary | 0.018 (0.513) | 0.018 (0.525) | 0.034 (0.356) | 0.041 (0.239) |
| Non-smokers with adenocarcinoma | 975/ 4415 | Primary | 0.012 (0.839) | 0.012 (0.829) | 0.054 (0.582) | 0.072 (0.387) |
|  |  | Secondary | 0.008 (0.916) | 0.053 (0.422) | 0.114 (0.182) | 0.103 (0.193) |
| Smokers with squamous cell | 4209/ 9084 | Primary | -0.019 (0.542) | -0.019 (0.554) | -0.084 (0.092) | 0.003 (0.938) |
|  |  | Secondary | -0.012 (0.679) | -0.012 (0.686) | -0.060 (0.146) | 0.002 (0.952) |
| Non-smokers with squamous cell | 158/ 4415 | Primary | -0.040 (0.773) | -0.040 (0.703) | 0.058 (0.777) | 0.034 (0.849) |
|  |  | Secondary | -0.217 (0.070) | -0.217 (0.069) | -0.342 (0.091) | -0.256 (0.130) |
| Smokers with small cell | 1761/ 9084 | Primary | -0.023 (0.593) | -0.023 (0.547) | 0.010 (0.884) | -0.003 (0.954) |
|  |  | Secondary | 0.001 (0.974) | 0.001 (0.967) | 0.027 (0.649) | -0.006 (0.910) |
| Non-smokers with small cell | 64/ 4415 | Primary | -0.383 (0.059) | -0.383 (0.052) | -0.329 (0.243) | -0.356 (0.170) |
|  |  | Secondary | -0.412 (0.021) | -0.412 (0.017) | -0.352 (0.163) | -0.361 (0.131) |

^1^The primary analysis included eight SNPs (rs729876, rs1537415, rs2738058, rs12461706, rs16870060, rs2521634, rs3826782, and rs7762544). The secondary analysis included six additional SNPs (rs1122900, rs2064712, rs2070901, rs4970469, rs9982623, and rs9984417).
^2^The primary Mendelian randomization methods was inverse‐variance weighted (IVW) MR. We used MR-PRESSO, simple median, and weighted median as secondary analyses. Betas indicate the effect estimate for the association between a one-unit increase in genetic predisposition to having chronic or aggressive periodontitis and the natural log risk for each outcome.

**Table S5.** Effect estimates for the association between genetic predisposition to having chronic or aggressive periodontitis and the risk of pancreatic cancer in the PanScan meta-analysis by sex, smoker status, study design, genetic instrument, and Mendelian randomization approach.

| Study design | Sub-set | N_cases_ / N_controls_ | Instrument^1^ | IVW^2^  β (p-value) | MR-PRESSO^2^ β (p-value) | Simple median β (p-value) | Weighted median β (p-value) |
| --- | --- | --- | --- | --- | --- | --- | --- |
| All | All | 5090/8733 | primary | 0.031 (0.372) | 0.031 (0.189) | 0.031 (0.537) | 0.026 (0.550) |
|  |  |  | secondary | 0.033 (0.262) | 0.033 (0.170) | 0.044 (0.312) | 0.026 (0.513) |
|  | Female | 2475/3048 | primary | 0.020 (0.691) | 0.020 (0.703) | -0.018 (0.803) | 0.017 (0.787) |
|  |  |  | secondary | 0.007 (0.877) | 0.007 (0.856) | -0.036 (0.567) | 0.015 (0.794) |
|  | Male | 2615/5685 | primary | 0.045 (0.347) | 0.045 (0.305) | 0.071 (0.318) | 0.011 (0.858) |
|  |  |  | secondary | 0.067 (0.107) | 0.067 (0.112) | 0.115 (0.077) | 0.014 (0.804) |
|  | Current smoker | 874/1286 | primary | 0.069 (0.437) | 0.069 (0.269) | 0.046 (0.720) | 0.031 (0.784) |
|  |  |  | secondary | 0.034 (0.664) | 0.034 (0.581) | 0.040 (0.725) | 0.030 (0.771) |
|  | Former smoker | 1871/3676 | primary | 0.053 (0.451) | 0.053 (0.476) | 0.033 (0.703) | 0.064 (0.399) |
|  |  |  | secondary | 0.074 (0.129) | 0.074 (0.149) | 0.071 (0.322) | 0.114 (0.086) |
|  | Never smoker | 1735/3388 | primary | -0.023 (0.692) | -0.023 (0.430) | 0.009 (0.909) | -0.017 (0.819) |
|  |  |  | secondary | -0.024 (0.631) | -0.024 (0.535) | 0.009 (0.898) | -0.019 (0.779) |
| Cohort | All | 1367/5120 | primary | 0.108 (0.117) | 0.108 (0.052) | 0.078 (0.430) | 0.100 (0.255) |
|  |  |  | secondary | 0.139 (0.020) | 0.139 (0.012) | 0.141 (0.114) | 0.124 (0.121) |
|  | Female | 705/1431 | primary | 0.123 (0.248) | 0.123 (0.286) | 0.128 (0.387) | 0.032 (0.805) |
|  |  |  | secondary | 0.156 (0.072) | 0.156 (0.080) | 0.164 (0.213) | 0.038 (0.749) |
|  | Male | 662/3689 | primary | 0.078 (0.425) | 0.078 (0.322) | 0.063 (0.654) | 0.159 (0.211) |
|  |  |  | secondary | 0.112 (0.187) | 0.112 (0.103) | 0.196 (0.116) | 0.193 (0.095) |
|  | Current smoker | 367/850 | primary | 0.087 (0.523) | 0.087 (0.468) | -0.097 (0.643) | 0.037 (0.838) |
|  |  |  | secondary | 0.127 (0.286) | 0.127 (0.210) | 0.021 (0.907) | 0.063 (0.698) |
|  | Former smoker | 485/2332 | primary | 0.128 (0.322) | 0.128 (0.355) | 0.057 (0.755) | 0.067 (0.670) |
|  |  |  | secondary | 0.173 (0.106) | 0.173 (0.130) | 0.037 (0.814) | 0.079 (0.574) |
|  | Never smoker | 461/1930 | primary | 0.036 (0.762) | 0.036 (0.768) | 0.109 (0.536) | -0.022 (0.891) |
|  |  |  | secondary | 0.075 (0.464) | 0.075 (0.471) | 0.130 (0.421) | -0.021 (0.887) |
| Case-control | All | 2537/2371 | primary | 0.039 (0.480) | 0.039 (0.263) | 0.078 (0.333) | 0.055 (0.437) |
|  |  |  | secondary | 0.006 (0.900) | 0.006 (0.870) | 0.076 (0.282) | 0.051 (0.418) |
|  | Female | 1148/956 | primary | 0.034 (0.679) | 0.034 (0.671) | -0.014 (0.915) | 0.047 (0.674) |
|  |  |  | secondary | -0.039 (0.582) | -0.039 (0.590) | -0.089 (0.428) | -0.056 (0.577) |
|  | Male | 1389/1415 | primary | 0.042 (0.567) | 0.042 (0.329) | 0.041 (0.689) | 0.037 (0.691) |
|  |  |  | secondary | 0.047 (0.459) | 0.047 (0.412) | 0.041 (0.653) | 0.035 (0.682) |
|  | Current smoker | 318/262 | primary | 0.125 (0.486) | 0.125 (0.477) | 0.244 (0.400) | 0.192 (0.446) |
|  |  |  | secondary | -0.005 (0.976) | -0.005 (0.976) | 0.128 (0.609) | 0.211 (0.350) |
|  | Former smoker | 1073/989 | primary | 0.065 (0.444) | 0.065 (0.433) | 0.060 (0.655) | 0.136 (0.211) |
|  |  |  | secondary | 0.068 (0.347) | 0.068 (0.223) | 0.108 (0.311) | 0.146 (0.127) |
|  | Never smoker | 903/1021 | primary | -0.010 (0.906) | -0.010 (0.854) | -0.017 (0.897) | -0.037 (0.733) |
|  |  |  | secondary | -0.061 (0.403) | -0.061 (0.278) | -0.077 (0.478) | -0.071 (0.473) |

^1^The primary analysis included eight SNPs (rs729876, rs1537415, rs2738058, rs12461706, rs16870060, rs2521634, rs3826782, and rs7762544). The secondary analysis included six additional SNPs (rs1122900, rs2064712, rs2070901, rs4970469, rs9982623, and rs9984417).
^2^The primary Mendelian randomization methods was inverse‐variance weighted (IVW) MR. We used MR-PRESSO, simple median, and weighted median as secondary analyses. Betas indicate the effect estimate for the association between a one-unit increase in genetic predisposition to having chronic or aggressive periodontitis and the natural log risk for each outcome.

**Table S6.** Effect estimates for the association between genetic predisposition to having chronic or aggressive periodontitis and the risk of pancreatic cancer in PanC4 by sex, smoker status, genetic instrument, and Mendelian randomization approach.

| Outcome | N_cases_ / N_controls_ | Instrument^1^ | IVW^2^  β (p-value) | MR-PRESSO β (p-value) | Simple median β (p-value) | Weighted median β (p-value) |
| --- | --- | --- | --- | --- | --- | --- |
| Overall | 4164/3792 | Primary | 0.000 (0.994) | 0.000 (0.992) | -0.020 (0.738) | -0.030 (0.548) |
|  |  | Secondary | 0.006 (0.855) | 0.006 (0.785) | 0.030 (0.551) | -0.042 (0.353) |
| Male | 2396/2106 | Primary | -0.045 (0.432) | -0.045 (0.457) | -0.028 (0.747) | -0.053 (0.448) |
|  |  | Secondary | -0.039 (0.388) | -0.039 (0.347) | -0.039 (0.574) | -0.056 (0.372) |
| Female | 1768/1686 | Primary | 0.057 (0.338) | 0.057 (0.305) | -0.022 (0.811) | 0.087 (0.278) |
|  |  | Secondary | 0.063 (0.218) | 0.063 (0.113) | 0.047 (0.539) | 0.118 (0.097) |
| Current smoker | 643/438 | Primary | 0.030 (0.783) | 0.030 (0.662) | -0.036 (0.814) | 0.032 (0.811) |
|  |  | Secondary | 0.020 (0.831) | 0.020 (0.793) | -0.036 (0.788) | -0.002 (0.985) |
| Former smoker | 1415/1306 | Primary | -0.093 (0.164) | -0.093 (0.075) | -0.099 (0.320) | -0.110 (0.208) |
|  |  | Secondary | -0.031 (0.590) | -0.031 (0.556) | -0.021 (0.822) | -0.099 (0.223) |
| Never smoker | 1579/1811 | Primary | 0.067 (0.310) | 0.067 (0.343) | 0.229 (0.046) | -0.008 (0.921) |
|  |  | Secondary | 0.058 (0.260) | 0.058 (0.232) | 0.057 (0.491) | 0.016 (0.828) |

^1^The primary analysis included eight SNPs (rs729876, rs1537415, rs2738058, rs12461706, rs16870060, rs2521634, rs3826782, and rs7762544). The secondary analysis included six additional SNPs (rs1122900, rs2064712, rs2070901, rs4970469, rs9982623, and rs9984417).
^2^The primary Mendelian randomization methods was inverse‐variance weighted (IVW) MR. We used MR-PRESSO, simple median, and weighted median as secondary analyses. Betas indicate the effect estimate for the association between a one-unit increase in genetic predisposition to having chronic or aggressive periodontitis and the natural log risk for each outcome.

**Funding and acknowledgments for the cancer consortia that provided genetic data for the analysis**

**Funding**

The pancreatic cancer consortia were supported by the NIH grants R21 CA234436, U19CA203654, and K12HD092535.

Genetics and Epidemiology of Colorectal Cancer Consortium (GECCO): National Cancer Institute, National Institutes of Health, U.S. Department of Health and Human Services (U01 CA164930, U01 CA137088, R01 CA059045, R01201407, R01CA189532).Genotyping/Sequencing services were provided by the Center for Inherited Disease Research (CIDR). CIDR is fully funded through a federal contract from the National Institutes of Health to The Johns Hopkins University, contract number HHSN268201200008I. This research was funded in part through the NIH/NCI Cancer Center Support Grant P30 CA015704.

ASTERISK: a Hospital Clinical Research Program (PHRC-BRD09/C) from the University Hospital Center of Nantes (CHU de Nantes) and supported by the Regional Council of Pays de la Loire, the Groupement des Entreprises Françaises dans la Lutte contre le Cancer (GEFLUC), the Association Anne de Bretagne Génétique and the Ligue Régionale Contre le Cancer (LRCC).

The ATBC Study is supported by the Intramural Research Program of the U.S. National Cancer Institute, National Institutes of Health, and by U.S. Public Health Service contract HHSN261201500005C from the National Cancer Institute, Department of Health and Human Services.

CLUE funding was from the National Cancer Institute (U01 CA86308, Early Detection Research Network; P30 CA006973), National Institute on Aging (U01 AG18033), and the American Institute for Cancer Research. The content of this publication does not necessarily reflect the views or policies of the Department of Health and Human Services, nor does mention of trade names, commercial products, or organizations imply endorsement by the US government.COLO2&3: National Institutes of Health (R01 CA60987).

ColoCare: This work was supported by the National Institutes of Health (grant numbers R01 CA189184 (Li/Ulrich), U01 CA206110 (Ulrich/Li/Siegel/Figueireido/Colditz, 2P30CA015704- 40 (Gilliland), R01 CA207371 (Ulrich/Li)), the Matthias Lackas-Foundation, the German Consortium for Translational Cancer Research, and the EU TRANSCAN initiative.

The Colon Cancer Family Registry (CCFR, www.coloncfr.org) is supported in part by funding from the National Cancer Institute (NCI), National Institutes of Health (NIH) (award U01 CA167551). The CCFR Set-1 (Illumina 1M/1M-Duo) and Set-2 (Illumina Omni1-Quad) scans were supported by NIH awards U01 CA122839 and R01 CA143247 (to GC). The CCFR Set-3 (Affymetrix Axiom CORECT Set array) was supported by NIH award U19 CA148107 and R01 CA81488 (to SBG). The CCFR Set-4 (Illumina OncoArray 600K SNP array) was supported by NIH award U19 CA148107 (to SBG) and by the Center for Inherited Disease Research (CIDR), which is funded by the NIH to the Johns Hopkins University, contract number HHSN268201200008I. The content of this manuscript does not necessarily reflect the views or policies of the NCI, NIH or any of the collaborating centers in the Colon Cancer Family Registry (CCFR), nor does mention of trade names, commercial products, or organizations imply endorsement by the US Government, any cancer registry, or the CCFR.

COLON: The COLON study is sponsored by Wereld Kanker Onderzoek Fonds, including funds from grant 2014/1179 as part of the World Cancer Research Fund International Regular Grant Programme, by Alpe d’Huzes and the Dutch Cancer Society (UM 2012–5653, UW 2013-5927, UW2015-7946), and by TRANSCAN (JTC2012-MetaboCCC, JTC2013-FOCUS). The Nqplus study is sponsored by a ZonMW investment grant (98-10030); by PREVIEW, the project PREVention of diabetes through lifestyle intervention and population studies in Europe and around the World (PREVIEW) project which received funding from the European Union Seventh Framework Programme (FP7/2007–2013) under grant no. 312057; by funds from TI Food and Nutrition (cardiovascular health theme), a public–private partnership on precompetitive research in food and nutrition; and by FOODBALL, the Food Biomarker Alliance, a project from JPI Healthy Diet for a Healthy Life.

Colorectal Cancer Transdisciplinary (CORECT) Study: The CORECT Study was supported by the National Cancer Institute, National Institutes of Health (NCI/NIH), U.S. Department of Health and Human Services (grant numbers U19 CA148107, R01 CA81488, P30 CA014089, R01 CA197350,; P01 CA196569; R01 CA201407) and National Institutes of Environmental Health Sciences, National Institutes of Health (grant number T32 ES013678).

CORSA: “Österreichische Nationalbank Jubiläumsfondsprojekt” (12511) and Austrian Research Funding Agency (FFG) grant 829675.

CPS-II: The American Cancer Society funds the creation, maintenance, and updating of the Cancer Prevention Study-II (CPS-II) cohort. This study was conducted with Institutional Review Board approval.

CRCGEN: Colorectal Cancer Genetics & Genomics, Spanish study was supported by Instituto de Salud Carlos III, co-funded by FEDER funds –a way to build Europe– (grants PI14-613 and PI09-1286), Agency for Management of University and Research Grants (AGAUR) of the Catalan Government (grant 2017SGR723), and Junta de Castilla y León (grant LE22A10-2). Sample collection of this work was supported by the Xarxa de Bancs de Tumors de Catalunya sponsored by Pla Director d’Oncología de Catalunya (XBTC), Plataforma Biobancos PT13/0010/0013 and ICOBIOBANC, sponsored by the Catalan Institute of Oncology.

Czech Republic CCS: This work was supported by the Grant Agency of the Czech Republic (grants CZ GA CR: GAP304/10/1286 and 1585) and by the Grant Agency of the Ministry of Health of the Czech Republic (grants AZV 15-27580A and AZV 17-30920A).

DACHS: This work was supported by the German Research Council (BR 1704/6-1, BR 1704/6-3, BR 1704/6-4, CH 117/1-1, HO 5117/2-1, HE 5998/2-1, KL 2354/3-1, RO 2270/8-1 and BR 1704/17-1), the Interdisciplinary Research Program of the National Center for Tumor Diseases (NCT), Germany, and the German Federal Ministry of Education and Research (01KH0404, 01ER0814, 01ER0815, 01ER1505A and 01ER1505B).

DALS: National Institutes of Health (R01 CA48998 to M. L. Slattery).

EDRN: This work is funded and supported by the NCI, EDRN Grant (U01 CA 84968-06).

EPIC: The coordination of EPIC is financially supported by International Agency for Research on

Cancer (IARC) and also by the Department of Epidemiology and Biostatistics, School of Public Health, Imperial College London which has additional infrastructure support provided by the NIHR Imperial Biomedical Research Centre (BRC).

The national cohorts are supported by: Danish Cancer Society (Denmark); Ligue Contre le Cancer, Institut Gustave Roussy, Mutuelle Générale de l’Education Nationale, Institut National de la Santé et de la Recherche Médicale (INSERM) (France); German Cancer Aid, German Cancer Research Center (DKFZ), German Institute of Human Nutrition Potsdam- Rehbruecke (DIfE), Federal Ministry of Education and Research (BMBF) (Germany); Associazione Italiana per la Ricerca sul Cancro-AIRC-Italy, Compagnia di SanPaolo and National Research Council (Italy); Dutch Ministry of Public Health, Welfare and Sports (VWS), Netherlands Cancer Registry (NKR), LK Research Funds, Dutch Prevention Funds, Dutch ZON (Zorg Onderzoek Nederland), World Cancer Research Fund (WCRF), Statistics Netherlands (The Netherlands); Health Research Fund (FIS) - Instituto de Salud Carlos III (ISCIII), Regional Governments of Andalucía, Asturias, Basque Country, Murcia and Navarra, and the Catalan Institute of Oncology - ICO (Spain); Swedish Cancer Society, Swedish Research Council and County Councils of Skåne and Västerbotten (Sweden); Cancer Research UK (14136 to EPIC-Norfolk; C8221/A29017 to EPIC-Oxford), Medical Research Council (1000143 to EPIC-Norfolk; MR/M012190/1 to EPIC-Oxford). (United Kingdom).

EPICOLON: This work was supported by grants from Fondo de Investigación Sanitaria/FEDER (PI08/0024, PI08/1276, PS09/02368, P111/00219, PI11/00681, PI14/00173, PI14/00230, PI17/00509, 17/00878, Acción Transversal de Cáncer), Xunta de Galicia (PGIDIT07PXIB9101209PR), Ministerio de Economia y Competitividad (SAF07-64873, SAF 2010-19273, SAF2014-54453R), Fundación Científica de la Asociación Española contra el Cáncer (GCB13131592CAST), Beca Grupo de Trabajo “Oncología” AEG (Asociación Española de Gastroenterología), Fundación Privada Olga Torres, FP7 CHIBCHA Consortium, Agència de Gestió d’Ajuts Universitaris i de Recerca (AGAUR, Generalitat de Catalunya, 2014SGR135, 2014SGR255, 2017SGR21, 2017SGR653), Catalan Tumour Bank Network (Pla Director d’Oncologia, Generalitat de Catalunya), PERIS (SLT002/16/00398, Generalitat de Catalunya), CERCA Programme (Generalitat de Catalunya) and COST Action BM1206 and CA17118. CIBERehd is funded by the Instituto de Salud Carlos III.

ESTHER/VERDI. This work was supported by grants from the Baden-Württemberg Ministry of Science, Research and Arts and the German Cancer Aid.

Harvard cohorts (HPFS, NHS, PHS): HPFS is supported by the National Institutes of Health (P01 CA055075, UM1 CA167552, U01 CA167552, R01 CA137178, R01 CA151993, R35 CA197735, K07 CA190673, and P50 CA127003), NHS by the National Institutes of Health (R01 CA137178, P01 CA087969, UM1 CA186107, R01 CA151993, R35 CA197735, K07CA190673, and P50 CA127003) and PHS by the National Institutes of Health (R01 CA042182).

Hawaii Adenoma Study: NCI grants R01 CA72520.

HCES-CRC: the Hwasun Cancer Epidemiology Study–Colon and Rectum Cancer (HCES-CRC; grants from Chonnam National University Hwasun Hospital, HCRI15011-1).

Kentucky: This work was supported by the following grant support: Clinical Investigator Award from Damon Runyon Cancer Research Foundation (CI-8); NCI R01CA136726.

LCCS: The Leeds Colorectal Cancer Study was funded by the Food Standards Agency and Cancer Research UK Programme Award (C588/A19167).

MCCS cohort recruitment was funded by VicHealth and Cancer Council Victoria. The MCCS was further supported by Australian NHMRC grants 509348, 209057, 251553 and 504711 and by infrastructure provided by Cancer Council Victoria. Cases and their vital status were ascertained through the Victorian Cancer Registry (VCR) and the Australian Institute of Health and Welfare (AIHW), including the National Death Index and the Australian Cancer Database.

MEC: National Institutes of Health (R37 CA54281, P01 CA033619, and R01 CA063464).

MECC: This work was supported by the National Institutes of Health, U.S. Department of Health and Human Services (R01 CA81488 to SBG and GR).

MSKCC: The work at Sloan Kettering in New York was supported by the Robert and Kate Niehaus Center for Inherited Cancer Genomics and the Romeo Milio Foundation. Moffitt: This work was supported by funding from the National Institutes of Health (grant numbers R01 CA189184, P30 CA076292), Florida Department of Health Bankhead-Coley Grant 09BN-13, and the University of South Florida Oehler Foundation. Moffitt contributions were supported in part by the Total Cancer Care Initiative, Collaborative Data Services Core, and Tissue Core at the H. Lee Moffitt Cancer Center & Research Institute, a National Cancer Institute-designated Comprehensive Cancer Center (grant number P30 CA076292).

NCCCS I & II: We acknowledge funding support for this project from the National Institutes of Health, R01 CA66635 and P30 DK034987.

NFCCR: This work was supported by an Interdisciplinary Health Research Team award from the Canadian Institutes of Health Research (CRT 43821); the National Institutes of Health, U.S. Department of Health and Human Serivces (U01 CA74783); and National Cancer Institute of Canada grants (18223 and 18226). The authors wish to acknowledge the contribution of Alexandre Belisle and the genotyping team of the McGill University and Génome Québec Innovation Centre, Montréal, Canada, for genotyping the Sequenom panel in the NFCCR samples. Funding was provided to Michael O. Woods by the Canadian Cancer Society Research Institute.

NSHDS: Swedish Research Council; Swedish Cancer Society; Cutting-Edge Research Grant and other grants from Region Västerbotten; Knut and Alice Wallenberg Foundation; Lion’s Cancer Research Foundation at Umeå University; the Cancer Research Foundation in Northern Sweden; and the Faculty of Medicine, Umeå University, Umeå, Sweden.

OFCCR: The Ontario Familial Colorectal Cancer Registry was supported in part by the National Cancer Institute (NCI) of the National Institutes of Health (NIH) under award U01 CA167551 and award U01/U24 CA074783 (to SG). Additional funding for the OFCCR and ARCTIC testing and genetic analysis was through and a Canadian Cancer Society CaRE (Cancer Risk Evaluation) program grant and Ontario Research Fund award GL201-043 (to BWZ), through the Canadian Institutes of Health Research award 112746 (to TJH), and through generous support from the Ontario Ministry of Research and Innovation.

OSUMC: OCCPI funding was provided by Pelotonia and HNPCC funding was provided by the NCI (CA16058 and CA67941).

PLCO: Intramural Research Program of the Division of Cancer Epidemiology and Genetics and supported by contracts from the Division of Cancer Prevention, National Cancer Institute, NIH, DHHS. Funding was provided by National Institutes of Health (NIH), Genes, Environment and Health Initiative (GEI) Z01 CP 010200, NIH U01 HG004446, and NIH GEI U01 HG 004438.

SCCFR: The Seattle Colon Cancer Family Registry was supported in part by the National Cancer Institute (NCI) of the National Institutes of Health (NIH) under awards U01 CA167551, U01 CA074794 (to JDP), and awards U24 CA074794 and R01 CA076366 (to PAN).

SEARCH: The University of Cambridge has received salary support in respect of PDPP from the NHS in the East of England through the Clinical Academic Reserve. Cancer Research UK (C490/A16561); the UK National Institute for Health Research Biomedical Research Centres at the University of Cambridge.

SELECT: Research reported in this publication was supported in part by the National Cancer Institute of the National Institutes of Health under Award Numbers U10 CA37429 (CD Blanke), and UM1 CA182883 (CM Tangen/IM Thompson). The content is solely the responsibility of the authors and does not necessarily represent the official views of the National Institutes of Health.

SMS and REACH: This work was supported by the National Cancer Institute (grant P01 CA074184 to J.D.P. and P.A.N., grants R01 CA097325, R03 CA153323, and K05 CA152715 to P.A.N., and the National Center for Advancing Translational Sciences at the National Institutes of Health (grant KL2 TR000421 to A.N.B.-H.)

The Swedish Low-risk Colorectal Cancer Study: The study was supported by grants from the Swedish research council; K2015-55X-22674-01-4, K2008-55X-20157-03-3, K2006-72X-20157-01-2 and the Stockholm County Council (ALF project).

Swedish Mammography Cohort and Cohort of Swedish Men: This work is supported by the Swedish Research Council /Infrastructure grant, the Swedish Cancer Foundation, and the Karolinska Institute´s Distinguished Professor Award to Alicja Wolk.

UK Biobank: This research has been conducted using the UK Biobank Resource under Application Number 8614

VITAL: National Institutes of Health (K05 CA154337).

WHI: The WHI program is funded by the National Heart, Lung, and Blood Institute, National Institutes of Health, U.S. Department of Health and Human Services through contracts HHSN268201100046C, HHSN268201100001C, HHSN268201100002C, HHSN268201100003C, HHSN268201100004C, and HHSN271201100004C.

**Acknowledgements:**

ASTERISK: We are very grateful to Dr. Bruno Buecher without whom this project would not have existed. We also thank all those who agreed to participate in this study, including the patients and the healthy control persons, as well as all the physicians, technicians and students.

CLUE: We appreciate the continued efforts of the staff members at the Johns Hopkins George W. Comstock Center for Public Health Research and Prevention in the conduct of the CLUE II study. We thank the participants in CLUE. Cancer incidence data for CLUE were provided by the Maryland Cancer Registry, Center for Cancer Surveillance and Control, Maryland Department of Health, 201 W. Preston Street, Room 400, Baltimore, MD 21201, http://phpa.dhmh.maryland.gov/cancer, 410-767-4055. We acknowledge the State of Maryland, the Maryland Cigarette Restitution Fund, and the National Program of Cancer Registries of the Centers for Disease Control and Prevention for the funds that support the collection and availability of the cancer registry data.

COLON and NQplus: the authors would like to thank the COLON and NQplus investigators at Wageningen University & Research and the involved clinicians in the participating hospitals.

CORSA: We kindly thank all those who contributed to the screening project Burgenland against CRC. Furthermore, we are grateful to Doris Mejri and Monika Hunjadi for laboratory assistance.

CPS-II: The authors thank the CPS-II participants and Study Management Group for their invaluable contributions to this research. The authors would also like to acknowledge the contribution to this study from central cancer registries supported through the Centers for Disease Control and Prevention National Program of Cancer Registries, and cancer registries supported by the National Cancer Institute Surveillance Epidemiology and End Results program.

Czech Republic CCS: We are thankful to all clinicians in major hospitals in the Czech Republic, without whom the study would not be practicable. We are also sincerely grateful to all patients participating in this study.

DACHS: We thank all participants and cooperating clinicians, and Ute Handte-Daub, Utz Benscheid, Muhabbet Celik and Ursula Eilber for excellent technical assistance.

EDRN: We acknowledge all the following contributors to the development of the resource: University of Pittsburgh School of Medicine, Department of Gastroenterology, Hepatology and Nutrition: Lynda Dzubinski; University of Pittsburgh School of Medicine, Department of Pathology: Michelle Bisceglia; and University of Pittsburgh School of Medicine, Department of Biomedical Informatics.

EPIC: Where authors are identified as personnel of the International Agency for Research on Cancer/World Health Organization, the authors alone are responsible for the views expressed in this article and they do not necessarily represent the decisions, policy or views of the International Agency for Research on Cancer/World Health Organization.

EPICOLON: We are sincerely grateful to all patients participating in this study who were recruited as part of the EPICOLON project. We acknowledge the Spanish National DNA Bank, Biobank of Hospital Clínic–IDIBAPS and Biobanco Vasco for the availability of the samples. The work was carried out (in part) at the Esther Koplowitz Centre, Barcelona.

Harvard cohorts (HPFS, NHS, PHS): The study protocol was approved by the institutional review boards of the Brigham and Women’s Hospital and Harvard T.H. Chan School of Public Health, and those of participating registries as required. We acknowledge Channing Division of Network Medicine, Department of Medicine, Brigham and Women's Hospital as home of the NHS. We would like to thank the participants and staff of the HPFS, NHS and PHS for their valuable contributions as well as the following state cancer registries for their help: AL, AZ, AR, CA, CO, CT, DE, FL, GA, ID, IL, IN, IA, KY, LA, ME, MD, MA, MI, NE, NH, NJ, NY, NC, ND, OH, OK, OR, PA, RI, SC, TN, TX, VA, WA, WY. The authors assume full responsibility for analyses and interpretation of these data.

Kentucky: We would like to acknowledge the staff at the Kentucky Cancer Registry.

LCCS: We acknowledge the contributions of Jennifer Barrett, Robin Waxman, Gillian Smith and Emma Northwood in conducting this study.

NCCCS I & II: We would like to thank the study participants, and the NC Colorectal Cancer Study staff.

NSHDS investigators thank the Biobank Research Unit at Umeå University, the Västerbotten Intervention Programme, the Northern Sweden MONICA study and Region Västerbotten for providing data and samples and acknowledge the contribution from Biobank Sweden, supported by the Swedish Research Council (VR 2017-00650).

PLCO: The authors thank the PLCO Cancer Screening Trial screening center investigators and the staff from Information Management Services Inc and Westat Inc. Most importantly, we thank the study participants for their contributions that made this study possible.

SCCFR: The authors would like to thank the study participants and staff of the Seattle Colon Cancer Family Registry and the Hormones and Colon Cancer study (CORE Studies).

SEARCH: We thank the SEARCH team.

SELECT: We thank the research and clinical staff at the sites that participated on SELECT study, without whom the trial would not have been successful. We are also grateful to the 35,533 dedicated men who participated in SELECT.

WHI: The authors thank the WHI investigators and staff for their dedication, and the study participants for making the program possible. A full listing of WHI investigators can be found at: <http://www.whi.org/researchers/Documents%20%20Write%20a%20Paper/WHI%20Investigator%20Short%20List.pdf>
